# Correlates of patient-reported cognitive performance with regard to disability

**DOI:** 10.1101/2021.02.02.21250685

**Authors:** Delphine Van Laethem, Alexander De Cock, Jeroen Van Schependom, Ralph HB Benedict, Guy Nagels, Marie D’hooghe

## Abstract

**Objectives:** The patient-reported Multiple Sclerosis Neuropsychological Questionnaire (MSNQ) is inconsistently related to objective cognitive tests in multiple sclerosis (MS), while being strongly correlated with depression. In this study we test whether the relation between subjective and objective cognitive performance is moderated by physical disability, assessed by the Expanded Disability Status Scale (EDSS).

**Materials & Methods:** From 275 MS patients who completed the patient-report MSNQ and the two-question screening tool for depression, we collected Symbol Digit Modalities Test (SDMT) and EDSS scores, indicators of respectively objective cognitive performance and physical disability. We analysed correlations between these variables in the total group and in three EDSS subgroups: Low 0.0 – 3.0, Medium 3.5 – 6.0 and High 6.5 – 9.0. We also investigated the use of a composite measure of cognitive impairment and depression.

**Results:** We found no significant correlations between patient-reported MSNQ and SDMT scores in the total group or the EDSS subgroups. MSNQ scores correlated significantly with depression in all subgroups. After correcting for several variables, MSNQ scores contributed adversely to SDMT scores in the total group, not in any subgroup. MSNQ scores contributed significantly to the prediction of the composite measure of impairment in the total group and in all EDSS subgroups.

**Conclusions:** The relationship between measures of subjective and objective cognitive performance is not influenced by the patient’s level of physical disability. MSNQ scores are substantially influenced by depression, and reflect cognitive function to some degree. The patient-report MSNQ can be useful to identify patients requiring further (neuro)psychological assessment.

## Introduction

Multiple sclerosis (MS), the most common inflammatory and neurodegenerative disease in young adults, affecting more than two million people worldwide^1^, is characterized by substantial clinical heterogeneity. Capturing the full spectrum of disease activity including the less visible burden remains a challenge^2^. Between 34 and 65 % of persons with multiple sclerosis (PwMS) suffer from cognitive impairment. Cognitive processing speed and visual learning and memory are impaired in more than 50% of PwMS, while verbal and working memory impairment is seen in about 30%. Impairment of visuospatial abilities and executive functions are found less commonly, in about 20% of patients^3-6^.

Because neuropsychological test batteries^7,8^ are time-consuming, single screening and monitoring tools are proposed. The Symbol Digit Modalities Test (SDMT), a sensitive measure of information processing speed^9^, is considered the best rapid assessment tool of cognition in clinical practice^10^ and has potential as a sentinel test for cognitive impairment in MS^11^. The Multiple Sclerosis Neuropsychological Questionnaire (MSNQ) was developed as a screening test for patients and informants to assess perceived cognitive impairment and to a lesser extent personality and behavioural changes without requiring professional expertise. The test consists of 15 questions that are scored by frequency of symptoms, with a patient-report and an informant-report version^12^.

However, the correlation patterns for the patient-report and informant-report version differ. While only the patient-report version correlated with screening measures for depression^12-16^, the correlations of the informant-report version with objective cognitive test results were consistently more pronounced than the correlations of the patient-report version with these tests^12,15-17^. Even though the patient-report version frequently but weakly correlated with objective cognitive test results, these findings resulted in the proposal to consider only the informant-report version as a sensitive and validated screening tool for cognitive impairment whereas increased MSNQ scores obtained by the patient were attributed to depression^13^. Benedict et al. proposed to use the patient-report MSNQ as a screening measure for cognitive impairment or depression, which nevertheless resulted in only a slight change in sensitivity, from 83% to 80%, and specificity, from 60% to 68%, when compared to just screening for cognitive impairment^17^. Recent findings of associations with reduced employment and work performance^18,19^, health-related quality of life^18^, health-promoting behaviours^18,20^ and reduced thalamic, cortical grey matter^21^ and hippocampal volumes^22^, nevertheless suggest a substantial impact of subjective cognitive complaints in PwMS, regardless of the link with depression.

An alternative explanation for the variable associations found between patient-reported and objective cognitive performance is the variation in EDSS scores. While Kurtzke Expanded Disability Status Scale (EDSS^23^) scores were not considered when selecting questions for the MSNQ^12^, later studies included patients with a wide variety of neurological disability^12-14,16,17^. We hypothesise that the patient-report MSNQ performs best as a cognitive screening measure in patients with lower EDSS scores, based on the assumption that several questions of the MSNQ may not apply to patients with higher EDSS scores. Increased physical disability is commonly associated with reduced activity and participation. Furthermore, these patients are at increased risk of failing objective cognitive tests^24,25^ even though they may not report cognitive symptoms^26,27^. Reduced awareness and lacking insight could explain why self-report MSNQ scores are not necessarily increased in a substantial proportion of patients with moderate to severe disability, despite objective cognitive impairment.

To evaluate our hypothesis, we investigate correlations between MSNQ and SDMT scores in low, medium and high EDSS groups with and without correcting for other variables. Furthermore, we assess the value of the patient-report MSNQ as a screening tool for both depression and cognitive impairment through the development of a composite measure of impairment in low, medium and high EDSS groups of a large patient sample.

## Materials & Methods

The study protocol, survey, patient information and informed consent were approved by the ethics committees at the University Hospital Brussels (B.U.N. 143201630261, 2016/357, 25 January 2017) and the National Multiple Sclerosis Center, Melsbroek (17/02, 24 January 2017). PwMS, aged 18 years or older, who were diagnosed with definite MS according to the McDonald criteria^28^ and registered in the EDMUS database from the University Hospital Brussels and the National Multiple Sclerosis Center, were included. 1908 patients received a postal survey, including the patient-report MSNQ and two-question screening tool for depression in MS, which is a reliable tool for identifying depression in PwMS, with a sensitivity of 98.5% and a specificity of 87%^29,30^. Patients were invited to contact a trained nurse by telephone in case of problems during completion of the questionnaire, to answer questions and administer the questionnaires together by telephone. Sex, age, onset date (date of first symptoms), phenotype at onset, years of education, SDMT score within the last six months before or after the survey (administered by trained nursing personnel) and EDSS score (assessed by trained neurologists) were retrieved from the database. For a more detailed description of the methods used, we refer to previous publications on this data set^16,18^. Since the scope of the study was to assess the value of the patient-reported MSNQ and the patients’ own assessment of cognition, the informant-report MSNQ was not included in the survey.

Patients were included if there was a full data set for the screening for depression, EDSS, MSNQ and SDMT scores. In case of an unanswered MSNQ question, this missing value was imputed by the mean item score. When more than one question of the MSNQ was not answered, this patient was excluded. The cohort was divided into three subcohorts according to EDSS scores: Low 0.0 – 3.0, Medium 3.5 – 6.0 and High 6.5 – 9.0. These cut-offs were based on the EDSS tertiles of our sample, as well as the severity of clinical and functional disease burden as estimated by experienced MS neurologists.

Results were reported as means with 95% confidence intervals (CI 95%), medians with ranges or as percentages. The relations between MSNQ, SDMT, EDSS and depression were investigated through Pearson correlations in the total population and EDSS subgroups. Multiple linear regression analyses were performed to estimate the contribution of age, depression and MSNQ to the SDMT score in the total population and EDSS subgroups. Furthermore, a composite measure of impairment was constructed, with patients being considered impaired if they were cognitively impaired, depressed or both. Cognitive impairment was defined as a normative SDMT-score (i.e. the raw SDMT is corrected based on the expected SDMT provided by a linear regression model trained on a group of healthy controls matched for age, years of education and sex^31^) of < -1.5 and depression was defined as answering ‘yes’ on at least one question of the two-question screening tool for depression in MS. A logistic regression analysis was performed to estimate the contribution of age, sex, years of education, EDSS and MSNQ to this composite measure. The performance of these models was assessed based on R^2^ and adjusted R^2^ and respectively t-tests and Wald tests were used to identify the significantly contributing explanatory variables. Finally, group differences in MSNQ scores between cognitively impaired versus cognitively preserved, depressed versus non-depressed and impaired versus non-impaired (according to the composite measure) patients were assessed visually through boxplots. A two-sided test with a type I error probability of 0.05 was used for all analyses.

## Results

### Population characteristics

277 patients met the inclusion criteria of our study. After visual inspection two patients with SDMT values of above 80 were removed from the data set, resulting in a total of 275 included patients. These SDMT outliers were patients who underwent monthly SDMT evaluations in the context of their treatment with natalizumab, which has led to an important learning effect due to repeated test exposure. In Table 1 the characteristics of the three EDSS subgroups and the total population are listed. The mean SDMT score was significantly different between the EDSS subgroups, with the low EDSS group scoring the highest and the high EDSS group the lowest. There was no significant difference in the mean MSNQ score between the three groups.

**Table 1:**
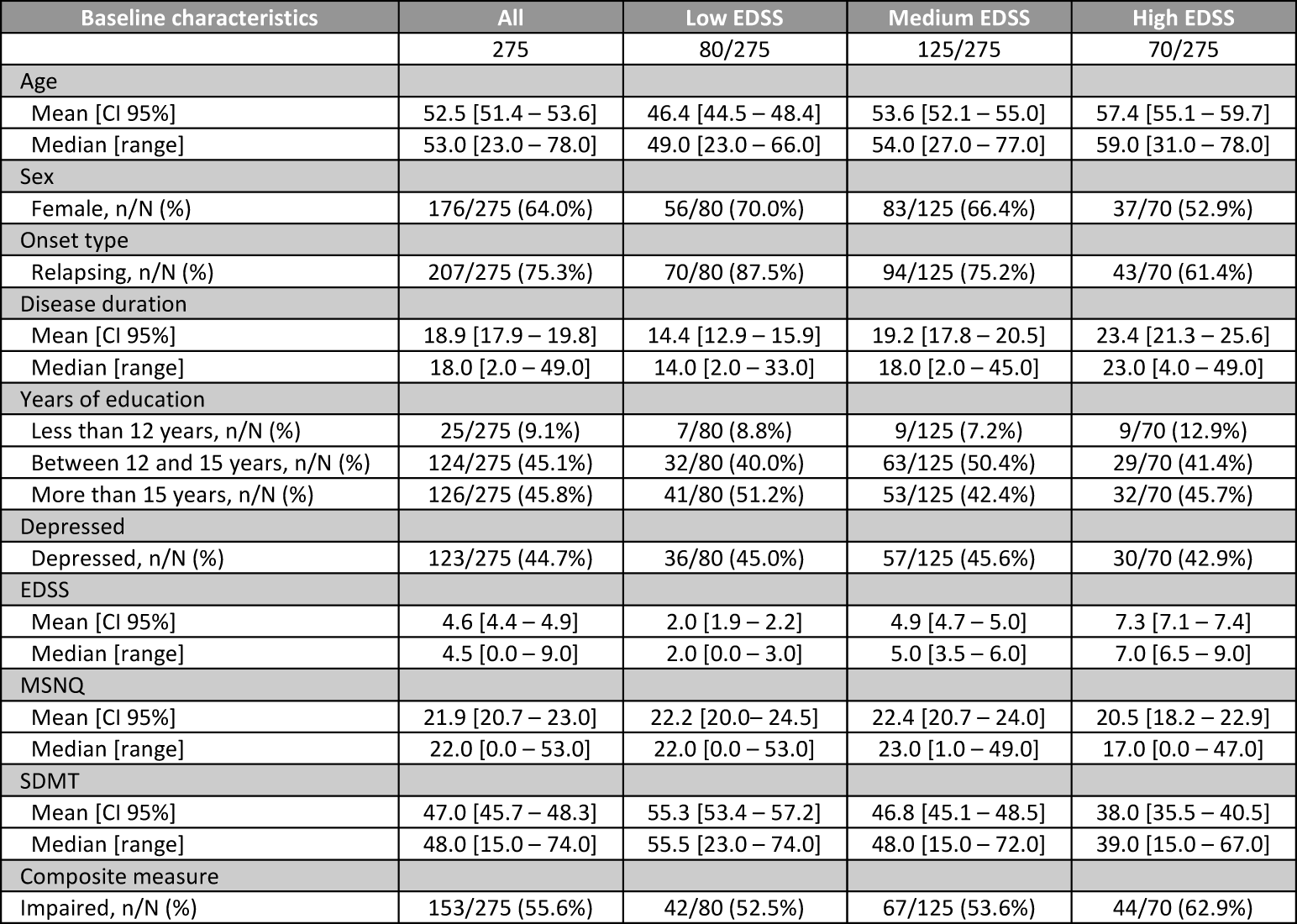
Population characteristics. CI 95% = 95% confidence interval, EDSS = Expanded Disability Status Scale, MSNQ = Multiple Sclerosis Neuropsychological Questionnaire, SDMT = Symbol Digit Modalities Test.

| Baseline characteristics | All | Low EDSS | Medium EDSS | High EDSS |
| --- | --- | --- | --- | --- |
|  | 275 | 80/275 | 125/275 | 70/275 |
| Age |  |  |  |  |
| Mean [CI 95%] | 52.5 [51.4 – 53.6] | 46.4 [44.5 – 48.4] | 53.6 [52.1 – 55.0] | 57.4 [55.1 – 59.7] |
| Median [range] | 53.0 [23.0 – 78.0] | 49.0 [23.0 – 66.0] | 54.0 [27.0 – 77.0] | 59.0 [31.0 – 78.0] |
| Sex |  |  |  |  |
| Female, n/N (%) | 176/275 (64.0%) | 56/80 (70.0%) | 83/125 (66.4%) | 37/70 (52.9%) |
| Onset type |  |  |  |  |
| Relapsing, n/N (%) | 207/275 (75.3%) | 70/80 (87.5%) | 94/125 (75.2%) | 43/70 (61.4%) |
| Disease duration |  |  |  |  |
| Mean [CI 95%] | 18.9 [17.9 – 19.8] | 14.4 [12.9 – 15.9] | 19.2 [17.8 – 20.5] | 23.4 [21.3 – 25.6] |
| Median [range] | 18.0 [2.0 – 49.0] | 14.0 [2.0 – 33.0] | 18.0 [2.0 – 45.0] | 23.0 [4.0 – 49.0] |
| Years of education |  |  |  |  |
| Less than 12 years, n/N (%) | 25/275 (9.1%) | 7/80 (8.8%) | 9/125 (7.2%) | 9/70 (12.9%) |
| Between 12 and 15 years, n/N (%) | 124/275 (45.1%) | 32/80 (40.0%) | 63/125 (50.4%) | 29/70 (41.4%) |
| More than 15 years, n/N (%) | 126/275 (45.8%) | 41/80 (51.2%) | 53/125 (42.4%) | 32/70 (45.7%) |
| Depressed |  |  |  |  |
| Depressed, n/N (%) | 123/275 (44.7%) | 36/80 (45.0%) | 57/125 (45.6%) | 30/70 (42.9%) |
| EDSS |  |  |  |  |
| Mean [CI 95%] | 4.6 [4.4 – 4.9] | 2.0 [1.9 – 2.2] | 4.9 [4.7 – 5.0] | 7.3 [7.1 – 7.4] |
| Median [range] | 4.5 [0.0 – 9.0] | 2.0 [0.0 – 3.0] | 5.0 [3.5 – 6.0] | 7.0 [6.5 – 9.0] |
| MSNQ |  |  |  |  |
| Mean [CI 95%] | 21.9 [20.7 – 23.0] | 22.2 [20.0 – 24.5] | 22.4 [20.7 – 24.0] | 20.5 [18.2 – 22.9] |
| Median [range] | 22.0 [0.0 – 53.0] | 22.0 [0.0 – 53.0] | 23.0 [1.0 – 49.0] | 17.0 [0.0 – 47.0] |
| SDMT |  |  |  |  |
| Mean [CI 95%] | 47.0 [45.7 – 48.3] | 55.3 [53.4 – 57.2] | 46.8 [45.1 – 48.5] | 38.0 [35.5 – 40.5] |
| Median [range] | 48.0 [15.0 – 74.0] | 55.5 [23.0 – 74.0] | 48.0 [15.0 – 72.0] | 39.0 [15.0 – 67.0] |
| Composite measure |  |  |  |  |
| Impaired, n/N (%) | 153/275 (55.6%) | 42/80 (52.5%) | 67/125 (53.6%) | 44/70 (62.9%) |

With regards to the composite measure of impairment, mean MSNQ scores were significantly different in depressed versus non-depressed and impaired versus non-impaired patients, but not in cognitively-impaired versus cognitively-preserved patients in the total population (see Figure 1) and all EDSS subgroups (not shown). The proportion of patients in the total population that is cognitively impaired versus preserved and depressed versus non-depressed is depicted in Figure 2.

**Figure 1:**
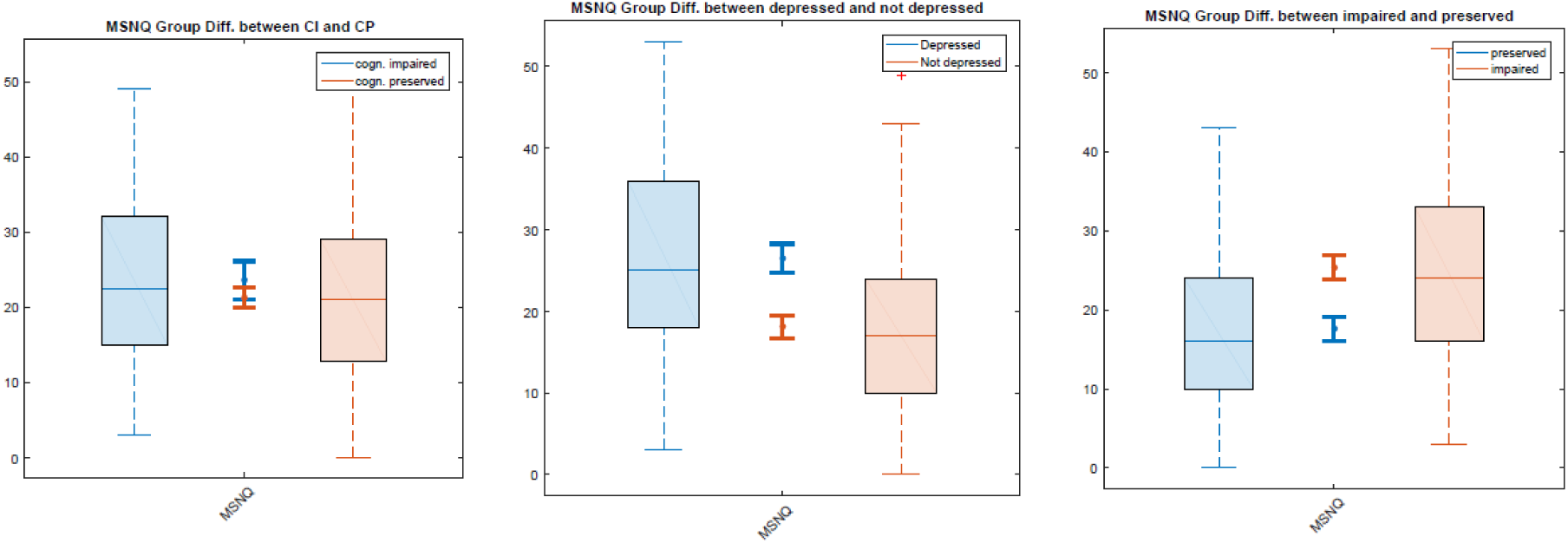
Group differences in MSNQ scores between cognitively impaired versus cognitively preserved, depressed versus non-depressed and impaired versus non-impaired patients. MSNQ = Multiple Sclerosis Neuropsychological Questionnaire, CI = cognitively impaired, CP = cognitively preserved, impaired = depression and/or cognitive impairment, preserved = no depression or cognitive impairment.

**Figure 2:**
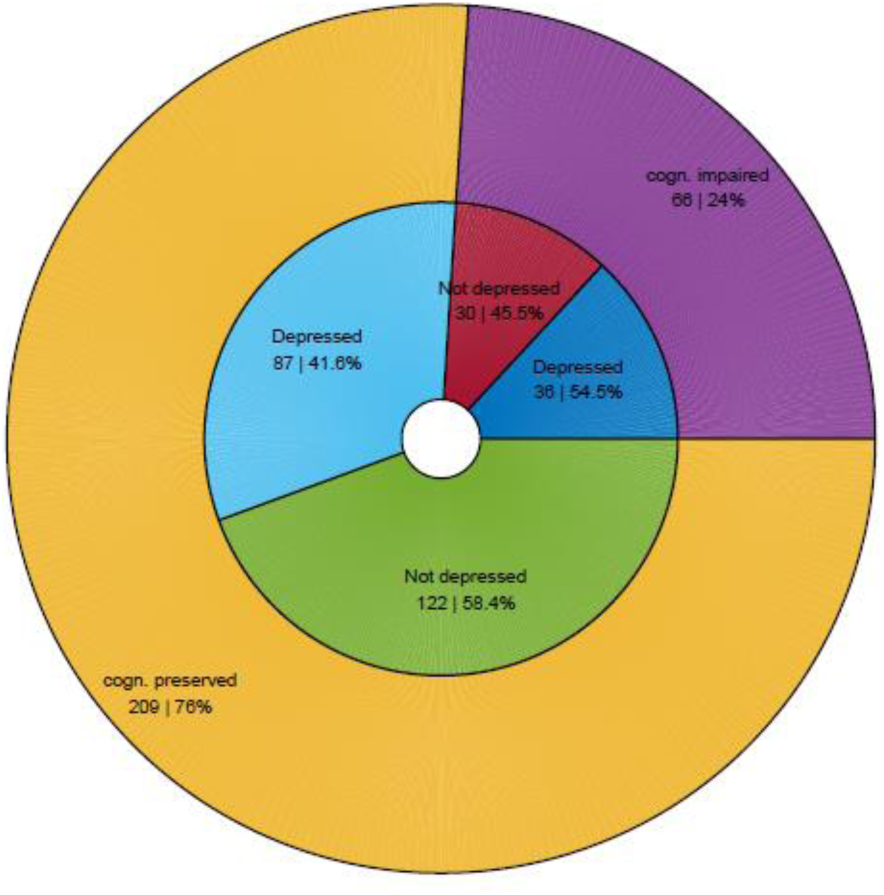
Composite measure of impairment in the total population.

### Correlations

As listed in Table 2, there were no significant correlations between MSNQ and SDMT scores in the total population or any of the EDSS subgroups. A positive correlation between EDSS and MSNQ scores was found in the low EDSS group only. MSNQ scores significantly correlated with depression in the total population and all EDSS subgroups and SDMT scores correlated with EDSS in the total population and the low and medium EDSS subgroup.

**Table 2:** Pearson correlations in the different EDSS subgroups. Correlations that are significant (*p* < 0.05) are marked with *. EDSS = Expanded Disability Status Scale, MSNQ = Multiple Sclerosis Neuropsychological Questionnaire, SDMT = Symbol Digit Modalities Test.

|  |  | All |  | Low EDSS |  | Medium EDSS |  | High EDSS |  |
| --- | --- | --- | --- | --- | --- | --- | --- | --- | --- |
| Var1 | Var2 | *r* | *p*-value | *r* | *p*-value | *r* | *p*-value | *r* | *p*-value |
| MSNQ | SDMT | -0.070 | 0.244 | -0.136 | 0.229 | -0.116 | 0.199 | -0.083 | 0.496 |
| EDSS | MSNQ | -0.067 | 0.271 | 0.233 | 0.038* | -0.138 | 0.124 | -0.193 | 0.110 |
| EDSS | SDMT | -0.544 | < 0.001* | -0.359 | 0.001* | -0.290 | 0.001* | -0.142 | 0.241 |
| MSNQ | Depression | 0.365 | < 0.001* | 0.372 | 0.001* | 0.371 | < 0.001* | 0.345 | 0.003* |

### Regression analyses

To estimate the combined contribution of age, depression and MSNQ to the SDMT score in the total population and the EDSS subgroups, multiple regression analyses were performed. Age, EDSS and MSNQ significantly and negatively contributed to the predicted value of the SDMT model of the total population (see Table 3). An increase of 1 year in age corresponded to a decrease of 0.20 points, an increase of 1 EDSS-point corresponded to a decrease of 2.93 points and an increase of 1 MSNQ-point corresponded to a decrease of 0.13 points in the predicted SDMT score respectively. None of the other variables significantly contributed. The contribution of MSNQ scores was no longer significant in the EDSS subgroup analyses.

**Table 3:** Multiple linear regression analysis of SDMT in the total population. Statistically significant predictors (p < 0.05) are marked with *. CI 95% = 95% confidence interval, EDSS = Expanded Disability Status Scale, MSNQ = Multiple Sclerosis Neuropsychological Questionnaire.

| N | R<sup>2</sup> | R<sup>2</sup><sub>adjusted</sub> | r |
| --- | --- | --- | --- |
| 275 | 0.33 | 0.32 | 0.580 |
| Variables | b (CI 95%) | b<sub>0</sub> (CI 95%) | p-value |
| Age | -0.20 ± 0.11 | -0.17 ± 0.09 | 0.002* |
| Sex: male | -0.15 ± 2.25 | -0.01 ± 0.17 | 0.913 |
| Years of education | 0.06 ± 0.31 | 0.02 ± 0.08 | 0.748 |
| EDSS | -2.92 ± 0.54 | -0.48 ± 0.09 | < 0.001* |
| Depression | -0.68 ± 2.31 | -0.05 ± 0.18 | 0.627 |
| MSNQ | -0.12 ± 0.10 | -0.11 ± 0.09 | < 0.05* |

To estimate the contribution of age, sex, years of education, EDSS and MSNQ to a composite measure of impairment (i.e. depression and/or cognitive impairment), a logistic regression analysis was performed. EDSS, years of education and MSNQ significantly contributed to the predicted value of the composite measure model of the total population (see Table 4). Furthermore, years of education was a positive predictor in the medium and high EDSS subgroups and MSNQ was a positive predictor in all subgroups.

**Table 4:** Logistic regression analysis of composite measure of impairment in the total population. Statistically significant predictors (p < 0.05) are marked with *. CI 95% = 95% confidence interval, EDSS = Expanded Disability Status Scale, MSNQ = Multiple Sclerosis Neuropsychological Questionnaire.

| N | R<sup>2</sup> | R<sup>2</sup><sub>adjusted</sub> |  |
| --- | --- | --- | --- |
| 275 | 0.15 | 0.13 |  |
| Variables | b (CI 95%) | b<sub>0</sub> (CI 95%) | p-value |
| Age | -0.00 ± 0.02 | -0.01 ± 0.24 | 0.943 |
| Sex: male | 0.27 ± 0.47 | 0.27 ± 0.47 | 0.347 |
| Years of education: | 0.14 ± 0.07 | 0.47 ± 0.23 | 0.001* |
| EDSS | 0.21 ± 0.12 | 0.45 ± 0.25 | 0.003* |
| MSNQ | 0.08 ± 0.02 | 0.93 ± 0.27 | < 0.001* |

## Discussion

We did not find significant correlations between MSNQ and SDMT scores in the total population or any of the EDSS subgroups. So, our hypothesis that the patient-report MSNQ performs better as a cognitive screening measure in patients with low compared to medium and high EDSS scores could not be confirmed. However, MSNQ scores contributed significantly and negatively to the prediction of SDMT scores after correcting for age and depression in the total population only, not in the EDSS subgroup analysis. These findings suggest that patient-reported MSNQ scores reflect cognitive functioning, as measured by the SDMT, at least to some degree. Furthermore, MSNQ scores contributed significantly to the prediction of a composite measure of cognitive impairment and depression after correcting for age, sex, years of education and EDSS, both in the total population and all EDSS subgroups.

Our findings confirm the association of patient-report MSNQ scores with depression across all EDSS subgroups. Furthermore, while MSNQ scores contributed significantly to the prediction of a composite measure of impairment (i.e. cognitive impairment and/or depression), this contribution was mainly driven by the association with depression. More specifically, mean MSNQ scores were significantly different in depressed versus non-depressed and impaired versus non-impaired patients (as assessed by our composite measure), but not in cognitively-impaired versus cognitively-preserved patients. The association of the patient-report MSNQ scores with depression is usually considered to be an important limitation when using it as a measure of cognition^12-16^. However, accurately measuring the cognitive issues caused by MS is challenging^32^ and the interplay between cognitive performance and depressive symptoms is complex. Cognitive impairment and depression are highly prevalent and underdiagnosed in PwMS^10,33^ and both have been associated with increased disability progression^34,35^. Depression also influences information processing speed, memory and executive function in PwMS and affects health behaviours and leisure activities, thereby reducing potential protective effects on cognition^36,37^. Previous studies with the Perceived Deficits Questionnaire have found evidence that subjective cognitive impairment reflects subtle declines in cognitive processing speed and memory independent of mood^27^ and that correlations between patient-reported memory and hippocampal volumes were maintained after controlling for depression^22^. While the patient-report MSNQ in our study was clearly influenced by depression, it also reflects a marker of cognitive function, as indicated by our multiple linear regression model of SDMT. These findings are in line with previously described results^14^. Therefore, reduced subjective cognitive performance and depressive symptoms should both be taken seriously and urge further investigation, especially when considering the important impact of these symptoms on daily life^18-20,33,35-37^. The patient-report MSNQ, which has been specifically developed for multiple sclerosis and is easy to administer without the need for professional expertise^12^, can therefore be a useful screening tool in daily practice for the identification of patients who are at risk of being depressed and/or cognitively impaired, and who thus require for further (neuro)psychological assessment. Furthermore, when used in combination with the informant-report version, the patient’s ability to estimate his own level over impairment can be assessed^26^.

Important strengths of our study are that, as far as we know, we are the first to assess the relationship between measures of objective and subjective cognitive performance in different EDSS groups. Furthermore, participants were recruited from two centres, representing a broad range of EDSS scores. A limitation is the cross-sectional study design. Moreover, as already discussed^16,18^, there was a moderate response rate to the survey, with a possibility of selection bias. Another weakness is that the MSNQ was sent by post and performed by the patient at home, which makes completion of the survey in a controlled environment impossible. Furthermore, the SDMT was assessed within six months before or after the survey, which could introduce bias since SDMT and MSNQ scores were not obtained at the same time. Another limitation is that only one objective cognitive test was carried out. Finally, it should be noted that when using a threshold of a normative SDMT of < -1.5 only 24% the included patients was cognitively impaired, which is lower than the expected 34 to 65%^3-6^. This could be due to the fact that patients were recruited either in an academic hospital (University Hospital Brussels) or a reference institute (National MS Center Melsbroek), where they regularly undergo cognitive testing, possibly resulting in a learning effect. The proportion of depressed patients in our study on the other hand corresponded to the expected prevalence of around 40%^33^.

In summary, we found that correlations between MSNQ and SDMT scores are not influenced by the patient’s level of physical disability. We were able to confirm patient-reported MSNQ scores are substantially influenced by depression, but nonetheless reflect cognitive impairment to some degree. Since both depression and cognitive impairment are underdiagnosed in PwMS and given the important impact of subjective cognitive symptoms on daily life, the patient-report MSNQ can be a useful screening tool for the identification of patients requiring further (neuro)psychological assessment and interventions.

## Data Availability

Data is available upon reasonable request to the authors.

## Acknowledgements

This research was funded by Biogen Fellowship Grant (ID #2016004-MS), which was provided by Biogen International GmbH to the National Multiple Sclerosis Center Melsbroek, Belgium (NMSC). Additional funding was provided by the MS Fonds KU Leuven. All patients who were willing to participate in our study are thanked for their time and effort.

This research was funded by Biogen Fellowship Grant (ID #2016004-MS), which was provided by Biogen International GmbH to the National Multiple Sclerosis Center Melsbroek, Belgium (NMSC). Additional funding was provided by the MS Fonds KU Leuven.

## References

1. Feigin VL, Krishnamurthi RV, Theadom AM, et al. Global, regional, and national burden of neurological disorders during 1990–2015: a systematic analysis for the Global Burden of Disease Study 2015. Lancet Neurol 2017;16(11):877–897.

2. van Munster CEP, Uitdehaag BMJ. Outcome Measures in Clinical Trials for Multiple Sclerosis. CNS Drugs 2017;31(3):217–236.

3. Amato MP, Zipoli V, Portaccio E. Multiple sclerosis-related cognitive changes: A review of crosssectional and longitudinal studies. J Neurol Sci 2006;245(1–2):41–46.

4. Bobholz J, Rao S. Cognitive dysfunction in multiple sclerosis: a review of recent developments. Curr Opin Neurol 2003;16:283–288.

5. Chiaravalloti ND, DeLuca J. Cognitive impairment in multiple sclerosis. Lancet Neurol 2008;7(12):1139–1151.

6. Benedict RHB, Amato MP, DeLuca J, et al. Cognitive impairment in multiple sclerosis: clinical management, MRI, and therapeutic avenues. Lancet Neurol 2020;19(10):860–871.

7. Sumowski JF, Benedict R, Enzinger C, et al. Cognition in multiple sclerosis: State of the field and priorities for the future. Neurology 2018;90(6):278–288.

8. Ruet A, Brochet B. Cognitive assessment in patients with multiple sclerosis: From neuropsychological batteries to ecological tools. Ann Phys Rehabil Med 2020;63(2):154–158.

9. Benedict RHB, Deluca J, Phillips G, et al. Validity of the Symbol Digit Modalities Test as a cognition performance outcome measure for multiple sclerosis. Mult Scler 2017;23(5):721–733.

10. Kalb R, Beier M, Benedict RHB, et al. Recommendations for cognitive screening and management in multiple sclerosis care. Mult Scler J 2018;24(13):1665–1680.

11. Van Schependom J, D’hooghe MB, Cleynhens K, et al. The Symbol Digit Modalities Test as sentinel test for cognitive impairment in multiple sclerosis. Eur J Neurol 2014;21(9):1219–e72.

12. Benedict RHB, Munschauer F, Linn R, et al. Screening for multiple sclerosis cognitive impairment using a self-administered 15-item questionnaire. Mult Scler 2003;9(1):95–101.

13. O’Brien A, Gaudino-Goering E, Shawaryn M, et al. Relationship of the Multiple Sclerosis Neuropsychological Questionnaire (MSNQ) to functional, emotional, and neuropsychological outcomes. Arch Clin Neuropsychol 2007;22(8):933–948.

14. Benedict RHB, Duquin JA, Jurgensen S, et al. Repeated assessment of neuropsychological deficits in multiple sclerosis using the symbol digit modalities test and the MS neuropsychological screening questionnaire. Mult Scler 2008;14(7):940–946.

15. Benedict RHB, Zivadinov R. Predicting neuropsychological abnormalities in multiple sclerosis. J Neurol Sci 2006;245(1–2):67–72.

16. B D’hooghe M, De Cock A, Van Remoortel A, et al. Correlations of health status indicators with perceived neuropsychological impairment and cognitive processing speed in multiple sclerosis. Mult Scler Relat Disord 2019;39:101904.

17. Benedict RHB, Cox D, Thompson LL, et al. Reliable screening for neuropsychological impairment in multiple sclerosis. Mult Scler 2004;10(6):675–678.

18. D’hooghe MB, De Cock A, Benedict RHB, et al. Perceived neuropsychological impairment inversely related to self-reported health and employment in multiple sclerosis. Eur J Neurol 2019;26(12):1447–1454.

19. Kobelt G, Langdon D, Jönsson L. The effect of self-assessed fatigue and subjective cognitive impairment on work capacity: The case of multiple sclerosis. Mult Scler J 2019;25(5):740–749.

20. Jelinek PL, Simpson S, Brown CR, et al. Self-reported cognitive function in a large international cohort of people with multiple sclerosis: associations with lifestyle and other factors. Eur J Neurol 2019;26(1):142–154.

21. Kletenik I, Alvarez E, Honce JM, et al. Subjective cognitive concern in multiple sclerosis is associated with reduced thalamic and cortical gray matter volumes. Mult Scler J Exp Transl Clin 2019;5(1):205521731982761.

22. Glukhovsky L, Brandstadter R, Leavitt VM, et al. Hippocampal volume is more related to patient-reported memory than objective memory performance in early multiple sclerosis. Mult Scler J 2021;27(4):568–578.

23. Kurtzke JF. Rating neurologic impairment in multiple sclerosis: An expanded disability status scale (EDSS). Neurology 1983;33(11):1444–1452.

24. Eizaguirre MB, Vanotti S, Merino Á, et al. The role of information processing speed in clinical and social support variables of patients with multiple sclerosis. J Clin Neurol 2018;14(4):472–477.

25. de Caneda MAG, de Vecino MCA. The correlation between EDSS and cognitive impairment in MS patients. Assessment of a Brazilian population using a BICAMS version. Arq Neuropsiquiatr 2016;74(12):974–981.

26. Carone DA, Benedict RHB, Munschauer FE, et al. Interpreting patient/informant discrepancies of reported cognitive symptoms in MS. J Int Neuropsychol Soc 2005;11(5):574–583.

27. Marrie RA, Chelune GJ, Miller DM, et al. Subjective cognitive complaints relate to mild impairment of cognition in multiple sclerosis. Mult Scler 2005;11(1):69–75.

28. Thompson AJ, Banwell BL, Barkhof F, et al. Diagnosis of multiple sclerosis: 2017 revisions of the McDonald criteria. Lancet Neurol 2018;17(2):162–173.

29. Mohr DC, Hart SL, Julian L, et al. Screening for depression among patients with multiple sclerosis: Two questions may be enough. Mult Scler 2007;13(2):215–219.

30. Minden SL, Feinstein A, Kalb RC, et al. Evidence-based guideline: Assessment and management of psychiatric disorders in individuals with MS Report of the Guideline Development Subcommittee of the American Academy of Neurology. Neurology 2014;82(2):174–181.

31. Costers L, Gielen J, Eelen PL, et al. Does including the full CVLT-II and BVMT-R improve BICAMS? Evidence from a Belgian (Dutch) validation study. Mult Scler Relat Disord 2017;18:33–40.

32. Leavitt, VM. The SDMT is not information processing speed. Mult Scler 2021;27(11): 1806–1807.

33. Marrie RA. Comorbidity in multiple sclerosis: Implications for patient care. Nat Rev Neurol 2017; 13(6):375–382.

34. Moccia M, Lanzillo R, Palladino R, et al. Cognitive impairment at diagnosis predicts 10-year multiple sclerosis progression. Mult Scler 2016;22(5):659–667.

35. Binzer S, McKay KA, Brenner P, et al. Disability worsening among persons with multiple sclerosis and depression: A Swedish cohort study. Neurology 2019;93(24):E2216–2223.

36. Patel VP, Walker LAS, Feinstein A. Revisiting cognitive reserve and cognition in multiple sclerosis: A closer look at depression. Mult Scler 2018;24(2):186–195.

37. Artemiadis A, Bakirtzis C, Ifantopoulou P, et al. The role of cognitive reserve in multiple sclerosis: A cross-sectional study in 526 patients. Mult Scler Relat Disord 2020;41:102047.

